# Decontaminating N95 respirators during the Covid-19 pandemic: simple and practical approaches to increase decontamination capacity, speed, safety and ease of use

**DOI:** 10.1101/2020.08.17.20177022

**Authors:** Riccardo Russo, Carly Levine, Courtney Grady, Blas Peixoto, Jessica McCormick-Ell, Thomas Block, Anthony Gresko, Guillaume Demas, Poonam Chitale, Alexis Frees, Alejandro Ruiz, David Alland

## Abstract

**Background:** The COVID-19 pandemic has caused a severe shortage of personal protective equipment (PPE), especially N95 respirators. Efficient, effective and economically feasible methods for large-scale PPE decontamination are urgently needed.

**Aims:** (1) to develop protocols for effectively decontaminating PPE using vaporized hydrogen peroxide (VHP); (2) to develop novel approaches that decrease set up and take down time while also increasing decontamination capacity (3) to test decontamination efficiency for N95 respirators heavily contaminated by makeup or moisturizers.

**Methods:** We converted a decommissioned Biosafety Level 3 laboratory into a facility that could be used to decontaminate N95 respirators. N95 respirators were hung on metal racks, stacked in piles, placed in paper bags or covered with makeup or moisturizer. A VHP^®^VICTORY^TM^ unit from STERIS was used to inject VHP into the facility. Biological and chemical indicators were used to validate the decontamination process.

**Findings:** N95 respirators individually hung on metal racks were successfully decontaminated using VHP. N95 respirators were also successfully decontaminated when placed in closed paper bags or if stacked in piles of up to six. Stacking reduced the time needed to arrange N95 respirators for decontamination by approximately two-thirds while almost tripling facility capacity. Makeup and moisturizer creams did not interfere with the decontamination process.

**Conclusions:** Respirator stacking can reduce the hands-on time and increase decontamination capacity. When personalization is needed, respirators can be decontaminated in labeled paper bags. Make up or moisturizers do not appear to interfere with VHP decontamination.

## Introduction

SARS-CoV-2, the virus responsible for the Covid-19 pandemic, has infected almost 17 million people and caused more than 680,000 deaths worldwide through July 2020 [1]. Healthcare workers (HCWs) can be exposed to high viral loads of SARS-CoV2 by inhaling droplets or aerosolized viral particles originating from patients under their care [2]. SARS-Cov-2 can survive in aerosols for more than 3 hours and the virus can be efficiently transmitted in this form [3,4]. In China, more than 3,000 HCWs were infected during the Covid-19 pandemic, and more than 10,600 workers were infected in Italy [5,6]. Shortages of personal protective equipment (PPE) can increase the risk of infection [2,7] and there is an increasing need to expand the manufacturing capability of PPE and to improve the supply chain [8]. Decontamination and reuse of PPE provides another means of increasing PPE availability. Several studies have validated the use of vaporized hydrogen peroxide (VHP) to decontaminate N95 respirators of bacteria, mycobacteria, viruses and importantly, SARS-CoV-2 [9,10]. An advantage of using VHP as a decontaminant is that it is degraded into oxygen and water eliminating concerns of toxic byproducts.

Here, we provide a detailed protocol for using VHP to effectively decontaminate N95 respirators and demonstrate that this protocol is effective even with N95 respirators that are heavily covered with makeup and moisturizers as might occur during day to day activities. Further, we describe and validate simple but novel modifications to this approach that can substantially increase the number of N95 respirators that can be decontaminated in a typical facility while simultaneously improving workflow and dramatically saving time and personnel efforts.

## Methods

### Operation Design

The Medical Science Building (MSB) located in the Newark campus of Rutgers University, holds a decommissioned Biosafety Level Three (BSL3) facility with a dedicated exhaust system making it possible to isolate and regulate airflow in this facility. This facility was also designed to maintain each room under negative pressure, in accordance with BSL3 design standards. These features enabled us to convert this facility into one that can be easily used to decontaminate N95 respirators. The facility has a 2,400 ft^3^ common room and three 1,200 ft^3^ modules. The common room was repurposed for N95 respirator decontamination and the three modules were partially sealed to contain VHP leakage. N95 respirators were hung on stainless racks using fifty metal hooks per shelf with five shelves per rack (Figure 1a). The hooks were secured to the racks using pliers. In other experiments N95 respirators were positioned in piles (Figures 1b and 1c) or inside paper bags (Figure 1d). To optimize exposure to VHP, N95 respirators were hung so that they did not touch each other. Seventeen metal racks were located along the walls of the common room leaving space available for additional racks if needed. Based on this design, the capacity for this operation was 4,250 N95 respirators per run using only the common room and the total capacity could be increased to 7,250 N95 respirators per run if racks were also located in the three modules.

**Figure 1:**
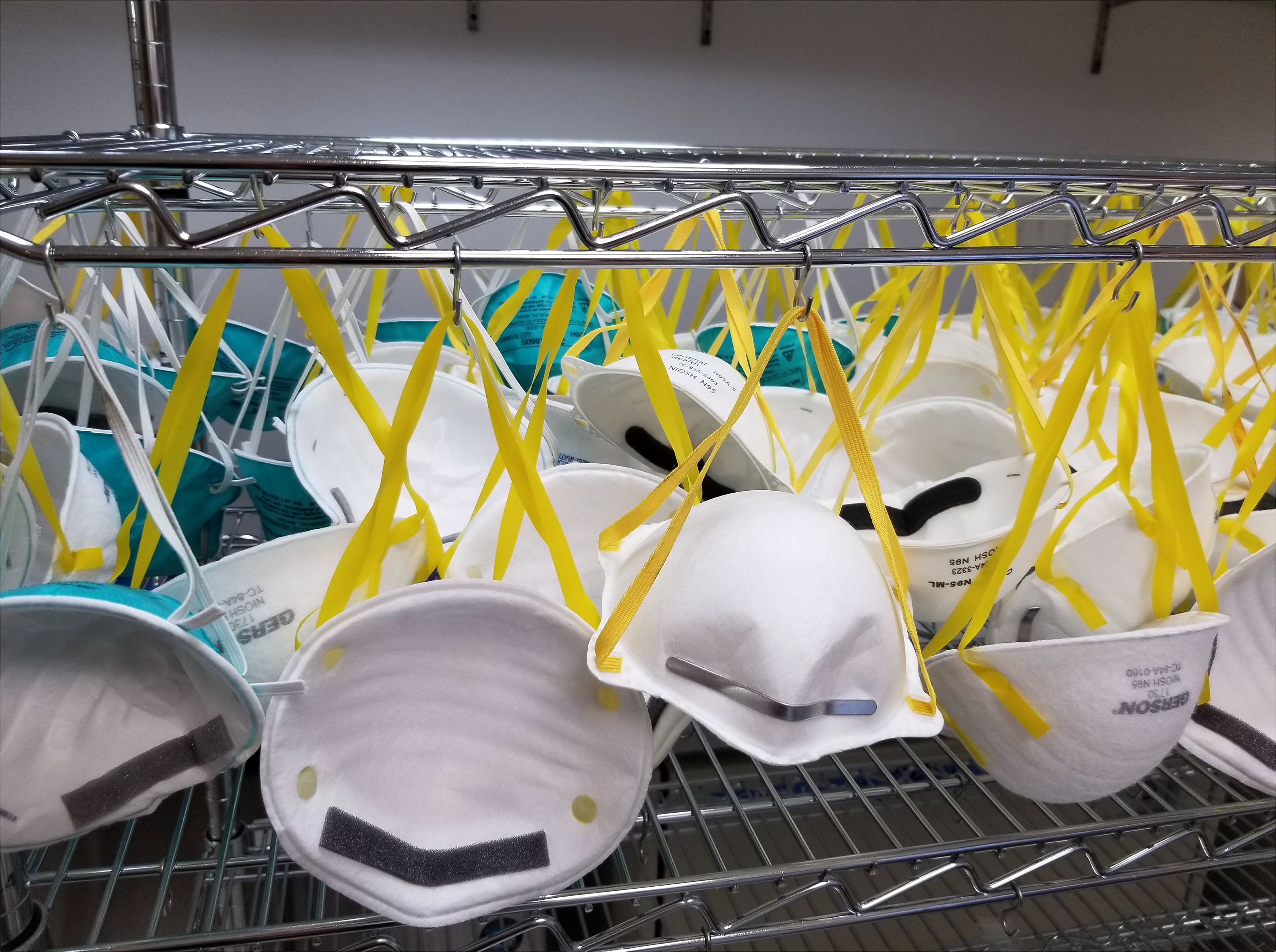
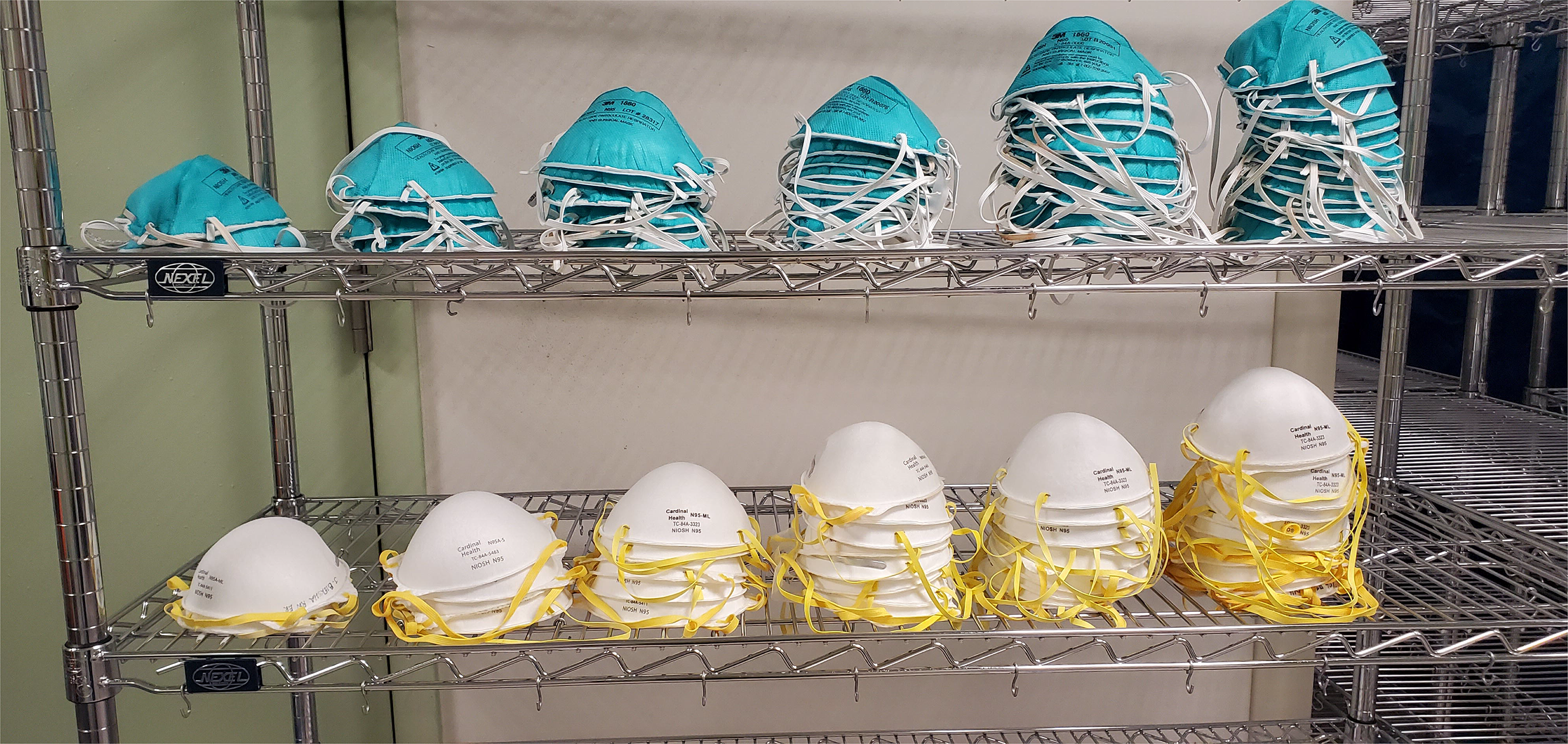
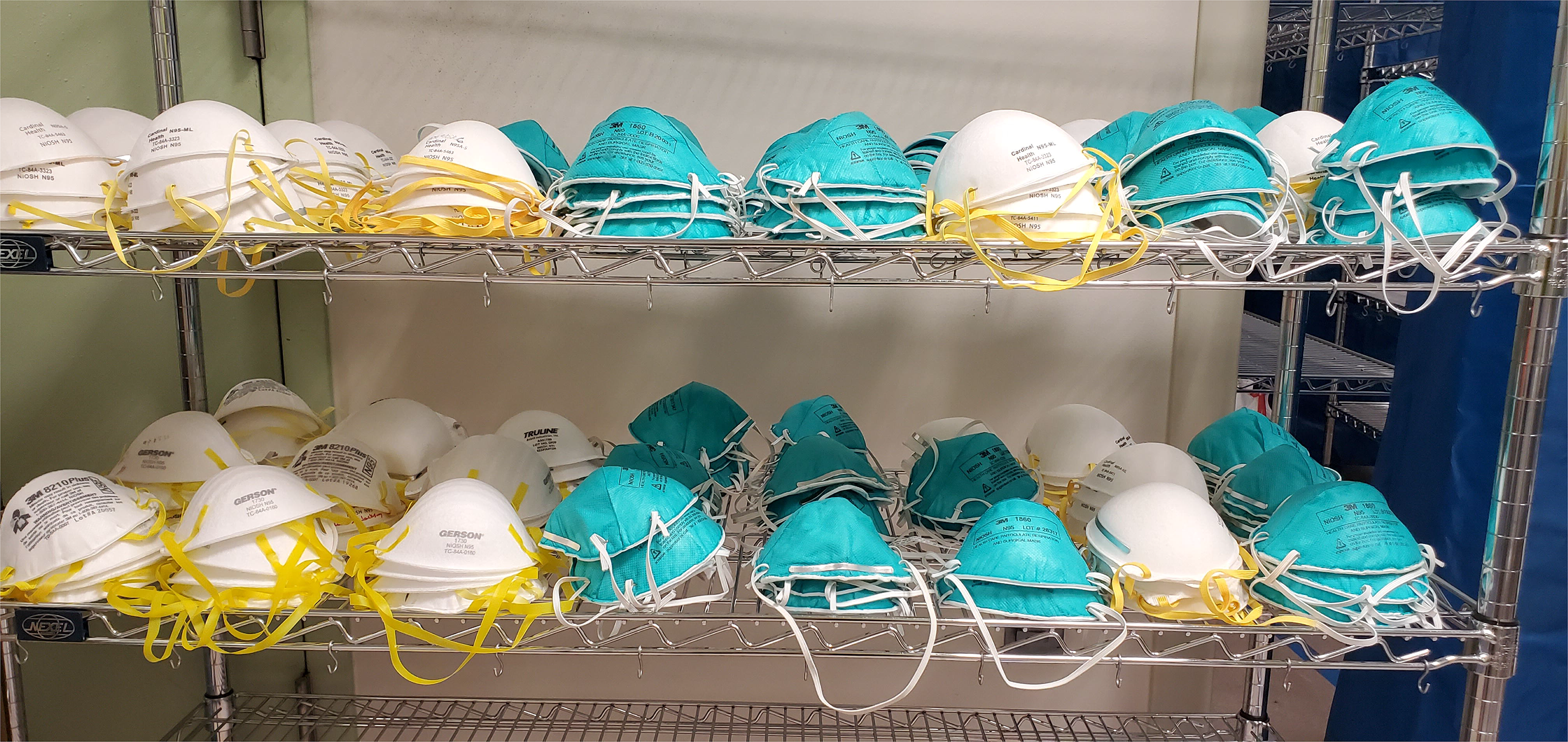
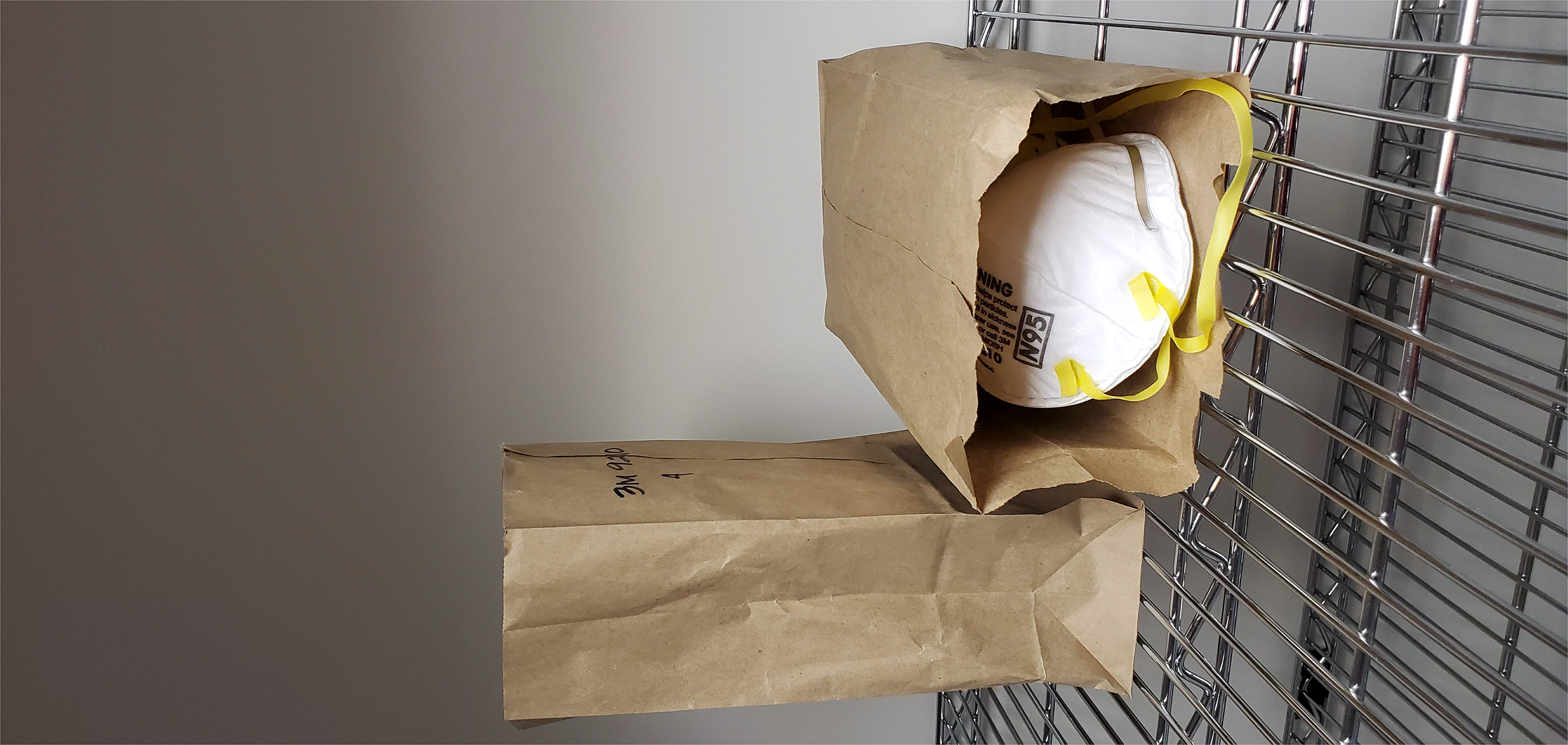
N95 respirator placement for decontamination of mobile racks. **A.** N95 respirators hung on the metal racks **B.** N95 respirator piles. Left to right stacks of 2, 4, 6, 8, 10 and 12 N95 respirators. **C** configuration of an entire rack of N95 respirators arranged in piles. **D.** N95 respirators individually packages in paper bags.

### Decontamination protocol

SARS CoV2 can survive prolonged periods on solid surfaces, and it is eliminated from cardboard after 24 hours [4]. As a result, multiple groups have recommended letting disposable N95 respirators ‘rest’ for at least 24 hours between uses [11]. All N95 respirators were kept in sealed boxes in a secure area for at least 48 hours before being hung for decontamination. Personnel wore PPE consisting of disposable Tyvek with booties and hoods, cover shoes, double gloves, aprons, P100 respirators and safety glasses to enter the facility and hang N95 respirators. It took a single individual an average of 30 minutes to hang 250 N95 respirators on one of our decontamination racks.

N95 respirators were then decontaminated with VHP using a VHP^®^ VICTORY^TM^ unit (Steris Life Sciences, Mentor, OH) filled with a 35% aqueous hydrogen peroxide solution (Vaprox^@^). The VICTORY^TM^ unit uses heat to vaporize hydrogen peroxide while maintaining the concentration below the dew point to keep hydrogen peroxide in a gaseous state thereby avoiding condensation. The concentration of VHP in the room was monitored using a VHP TS1000 Tri-Scale Sensor. If needed, a VHP AR12000 Aerator could be used to remove VHP from the room. The VICTORY^TM^ unit was remotely controlled by a computer using *SmartPhase*^TM^ software technology that automatically adjusted VHP injection rates based on room temperature, relative humidity, and VHP concentration. The program automatically calculated bioburden reduction and VHP saturation in real-time to maintain the concentration of VHP close to the setting point and to maintain the vapor state, respectfully.

Four fans were positioned in different areas of the common room to ensure a uniform distribution of VHP. Since VHP is more dense than air, fans were oriented to blow air toward the ceiling to facilitate the dispersion of VHP in the higher regions. The locations of the fans were marked on the ground using masking tape to ensure that they maintained positions during each decontamination run.

Decontamination was performed in four phases: conditioning, gassing, gassing-dwell and aeration. Based on the temperature, humidity and volume of the facility, the condition and gassing phases lasted approximately 90 minutes. During these phases, VHP was injected at a rate between 5 to 40 g/min to keep a target concentration of 400 ppm. The actual concentration varied between 400 and 800 ppm. The concentration of VHP was remotely monitored using the VNC Viewer software on a portable computer. The dwelling phase was maintained for 3 hours with no additional VHP injected during this time. Supply and exhaust fans were switched back on at the end of the dwelling phase to facilitate the removal of the VHP. The aeriation phase was kept overnight for approximately 15-18 hours. The following morning, the residual concentration of VHP in the facility was measured using a Dräger x-Am 5100 (Dräger, Telford, PA) to ensure that the VHP concentration was below the safe level of 1 ppm.

Five validation runs were conducted on approximately 1,250 N95 respirators of different models. A rack with 250 control N95 respirators was decontaminated with every cycle to determine the effect of multiple sterilization cycles on the integrity of the N95 respirators. Biological indicators (BIs) (Spordex^®^ VHP Biological Indicator Discs, Steris Life Sciences, Mentor, OH) and chemical indicators (CIs) (Steraffin^®^ VHP Type 4 Process Indicator, Steris Life Sciences, Mentor, OH) were positioned in different parts of the room. Some indicators were also placed within closed control N95 respirators to validate that the all areas were exposed to the correct concentration of VHP. After decontamination, CIs were inspected visually to observe if it had changed from violet to yellow indicating that VHP had successfully contacted the area. BIs containing *Geobacillus stearothermophilus* spores were aseptically transferred into Tryptic Soy Broth and incubated for 7 days at 37 C. Results were read at 24 hours and 7 days post inoculation. Positive and negative controls were included in every run to represent BI viability and media sterility.

### N95 respirators

Respirator models 3M 9210, 3M 1870, 3M 1870+, 3M 1860S, and 3M 1860 (3M, St. Paul, MN; Cardinal Health, Dublin, OH, Gerson, Middleboro, MA); Cardinal Health S and M/L (Cardinal Health, Dublin, OH), Gerson 2130 and Gerson 1730; (Gerson, Middleboro, MA), Halyard Fluidshield 46727 and 46827; (O&M Halyard, Inc., Alpharetta, GA) were tested in this study. These are the models most frequently used by HCWs at University Hospital (UH, Newark NJ).

### Quality control

At the end of each decontamination cycle, the N95 respirators were inspected to verify that there was no physical damage or loss of integrity. N95 respirators in the control rack underwent twelve decontamination cycles and were inspected after each run to determine if there was loss in respirator integrity, the results of the respirator integrity checks are described in a separate study submitted at bioRxiv (Levine et al.).

### Improving workflow using N95 respirator piles

Trials were conducted to identify ways to increase the number of N95 respirators that could be decontaminated at the same time in our facility by replacing the tedious N95 respirator hanging process with a faster method. For this purpose, five N95 models (Table I) were piled in stacks of four, six, eight, ten and twelve units (Figure 1b). BIs and CIs were inserted into the middle of each pile to determine which pile configuration remained consistent with full N95 respirator decontamination.

**Table I:** Decontamination results when N95 respirators were placed in piles of various sizes.

| **Respirator Model** | **Number of respirators per pile** | **n. of replicates** | **n. of experiments** | **BIs and CIs results** |
| --- | --- | --- | --- | --- |
| Gerson 1730<br>(TC-84A-0160) | 4 | 3 | 4 | pass |
|  | 6 | 3 | 4 | pass |
|  | 8 | 1 | 1 | pass |
|  | 10 | 1 | 1 | pass |
| 3M 1860S (TC-84A-0006) | 4 | 3 | 4 | pass |
|  | 6 | 3 | 4 | pass |
|  | 8 | 1 | 1 | pass |
|  | 10 | 1 | 1 | failed |
|  | 12 | 1 | 1 | failed |
| 3M 8210 Plus | 4 | 3 | 4 | pass |
|  | 6 | 3 | 3 | pass |
| Truline A501780 | 4 | 3 | 4 | pass |
|  | 6 | 3 | 4 | pass |
|  | 8 | 1 | 1 | pass |
|  | 10 | 1 | 1 | pass |
| Cardinal Health<br>N95A-S | 4 | 3 | 4 | pass |
|  | 6 | 3 | 4 | pass |
|  | 8 | 1 | 2 | pass |
|  | 10 | 1 | 2 | pass |
|  | 12 | 1 | 2 | pass |

### Improving workflow by decontaminating N95 respirators in paper bags

HCWs at UH collected their N95 respirators in paper bags. Each bag was labelled with the user’s name and contained either one or two N95 respirators. When the N95 respirators were received at the decontamination site, they were removed from the bag and hung on metal racks. This made it difficult to return N95 respirators to their original owners. Decontaminating N95 respirators without removing them from the paper bags could improve workflow and simplify the process needed to return N95 respirators to their original users. To test the safety of this procedure modification, a sample of five N95 respirators (Halyard Fluidshield 46727/46827) were put inside five different paper bags (Figure 1d), the N95 respirators were closed with tape to mimic a worst case scenario where a N95 respirator was completely folded, and the bag was closed (in a normal operation the bags would be left open to facilitate the dispersion of VHP). BIs and CIs were inserted inside closed N95 respirators within the paper bags. The trial was repeated in five different decontamination runs.

### Effect of makeup and moisturizer creams on decontamination efficacy

Several N95 respirators received from UH for decontamination had extensive traces of makeup or moisturizer creams. To determine if a layer of makeup or moisturizer cream could interfere with the decontamination process, N95 respirators (Halyard Fluidshield 46727/46827) were covered with makeup (Maybelline Fit Me Matte + Poreless Foundation, L’Oreal, New York, NY), lip stick (Maybelline Lipstick, L’Oreal, New York, or moisturizer cream (Oil Free Moisturizer, Beauty 360, CVS Woonsocket, RI) or a combination of makeup and moisturizer cream, BIs and CIs were placed inside the N95 respirator, and the N95 respirator was then folded and closed using tape. Halyard Fluidshield (model 46727/46827) N95 respirators were selected for this study since they can be folded. The trial was repeated in five different decontamination runs.

## Results

### Facility Decontamination

An example of the decontamination cycle is reported in the supplement Figure S2. On average, it took 90 minutes for VHP concentrations to reach a level reported to reduce microorganisms by 12-logs based on calculations from the *SmartPhase*^TM^ software. The concentration of VHP fell below 1 ppm approximately 5 to 6 hours after turning on supply and exhaust fans. Visual inspection of CIs after the decontamination cycle and the BIs 7 days post inoculation, validated successful decontamination at all locations within the facility. Additionally, CIs located inside control N95 respirators turned yellow and all BIs were negative for growth after 7 days post inoculation, indicating that all surfaces of the N95 respirators were decontaminated even when folded.

### Improving workflow using N95 respirator piles

One to four piles containing stacks of four, six, eight, ten or twelve N95 respirators per pile were decontaminated for each of five N95 respirator models (Table I). The results of the BIs and CIs that had been inserted into the middle of the piles were used to determine whether effective decontamination had occurred. Our results showed that all N95 respirator models were decontaminated when stacked in piles of eight or fewer. However, the 3M 1860S (TC-84A-0006) N95 respirators were not decontaminated when piled in stacks of ten or twelve. All other N95 respirator models were decontaminated even when piled in stacks of ten or twelve. Four additional decontamination experiments were then performed on piles of four and six N95 respirators using all five N95 respirator models. In these follow up experiments, both four and six N95 respirator piles and all five models were decontaminated according to both the BI and CI indicators.

On each shelf we were able to place 24 piles of six N95 respirators (Figure 1c), for a total of 720 units per rack. When we hung N95 respirators, we were able to place 50 units per shelf, for a total of 250 per rack. In our operation we used 17 racks. Based on this design, we would have been able to increase our decontamination capacity from 4,250 to 12,240 units per run when N95 respirators were arranged in piles instead of hanging them.

Placing the N95 respirators in piles also saved time in the setting up and taking down process. It took approximately 8 minutes to fill a rack with piles of N95 respirators, compared to 30 minutes when they were hung. Based on our operation design, it took in total 2 hours and 20 minutes to arrange the N95 respirators in the 17 racks compared to 8 hours and 30 minutes when they were hung. It took a similar time to remove the N95 respirators from the shelves after the decontamination.

### N95 respirators in paper bags

Three replicates of N95 respirators folded up inside paper bags were tested in five separate experiments. BIs and CIs inserted in folded and closed N95 respirators inside the closed paper bags were used to test for decontamination. Our results showed that effective decontamination for all N95 respirators in all experiments (Table II).

**Table II:** Decontamination results when N95 respirators were covered with makeup and moisturize cream or placed inside paper bags.

| **Respirator Model** | **Condition** | **n. of replicates** | **n. of experiments** | **BIs and CIs results** |
| --- | --- | --- | --- | --- |
| Halyard<br>Fluidshield<br>46727/46827 | Respirator covered in moisturizer cream | 3 | 5 | pass |
|  | Respirator covered in foundation | 3 | 5 | pass |
|  | Respirator covered in moisturizer cream and foundation | 3 | 5 | pass |
| Halyard<br>Fluidshield<br>46727/46827 | Inside brown paper bag | 3 | 5 | pass |

### Makeup and moisturizer creams

Halyard Fluidshield (model 46727/46827) N95 respirators were used to assess whether makeup and/or moisturizer creams interfere with the ability of VHP to decontaminate. BIs and Cis placed inside the N95 respirators were used to test for decontamination. Neither makeup nor moisturizer cream, nor the combination of both interfered with the decontaminated procedure, across five separate experiments (Table II).

## Discussion

We have described how to decontaminate thousands of N95 respirators using a relatively small 2,400 ft^3^ facility. The entire decontamination process, from hanging the N95 respirators to the end of the sterilization cycle, took an entire day using a conventional mask hanging protocol. We hung N95 respirators as the first task of the morning and then started the sterilization cycle later in the morning. The aeration cycle to remove VHP from the decontamination facility was then started in the late afternoon, and we went inside the facility the following morning when the VHP concentration was close to 0 ppm. Toward the end of the project, we hung N95 respirators the day before running a decontamination cycle to facilitate the process. Since the N95 respirators were kept in a BSL3 facility that is under negative pressure and locked, there was no risk of exposure to people outside the facility. When we ran decontamination cycles daily, we observed that the concentration of VHP would not fall below 1 ppm on the following day. It is possible that the walls, ceiling and floors of the facility became saturated with VHP when the decontamination cycle was run daily since the situation did not change even when there were less than one hundred N95 respirators in the facility. Thus, we recommend every other day sterilization in facilities where this problem occurs. Some HCWs wear makeup or use moisturizer creams that can leave traces on N95 respirators, even ones destined for decontamination and reuse. Here we demonstrated that makeup or moisturizer creams are unlikely to interfere with the decontamination process when VHP is used. Our study describes a more efficient way to perform the decontamination process. We have also confirmed that most but not all models of N95 respirators will continue to fit and function properly for at least six decontamination cycles in a separate publication submitted at bioRxiv (Levine et al.). This has also been confirmed by others using a single N95 respirator model [12].

Two of the goals of our study were to find ways simplify the process of preparing N95 respirators for decontamination and to increase the overall number of N95 respirators that could be decontaminated in a single day. We tested the possibility of piling N95 respirators on top of each other in small stacks instead of hanging them. In these trials, we tested respirators from several brands to determine if this process could be valid for different N95 respirator models. For one model, piles of ten or twelve N95 respirators were not always completely decontaminated. However, we observed in repeated tests that all respirator models could be decontaminated in piles of four, six or eight. Based on these results, it seems likely that N95 respirators can be consistently decontaminated in pile sizes of up to six units, but each facility should validate this process.

Piling N95 respirators will speed up the preparation process and significantly increase the capacity of a decontamination facility. It took 30 minutes for a single individual to hang 250 N95 respirators on one rack. In contrast, it took 8 minutes to pile the same number of N95 respirators on a rack. This means that it would take 8.5 hours for a single individual to hang 4,250 N95 respirators compared 2.5 hours if N95 respirators were piled as we have described. Furthermore, piles take up substantially less space than hung N95 respirators. It follows that using a six-stack piling strategy would increase the capacity of a facility such as ours from 4,250 to 12,240 units. Piling N95 respirators would also enable all hooks to be eliminated from the decontamination racks. Hooks can tear the PPE of personal hanging respirators; thus, eliminating hooks would improve the overall safety of the procedure.

We also looked at ways to improve the sorting process of N95 respirators after decontamination. Our experience is that HCWs prefer to receive their own originally worn N95 respirators put in paper bags marked with their name. For this reason, we decontaminated N95 respirators kept in paper bags, but we replicated the worse possible scenario sealing the bags and inserting CIs and BIs inside N95 respirators that were closed. We found that it is possible to decontaminate N95 respirators under the conditions tested. Leaving N95 respirators in paper bags would also speed up and facilitate the sorting of N95 respirators after the decontamination process and will not significantly decrease the capacity of the facility. We estimated that we could hang 50 N95 respirators per shelf or put up to 20 paper bags containing one or two N95 respirator per shelf.

## Conclusion

In this project, we were able to design an inexpensive reproducible operation for the decontamination of N95 respirators using VHP. Some advantages of this operation were that it required a small facility and that the N95 respirator capacity could be easily increased by our modified procedures. We also reported possible ways to speed up the process of preparing N95 respirators for decontamination by arranging them in piles of four or six instead on hanging. Finally, we validated the potential to keep N95 respirators as they were received, in paper bags, during the decontamination process thus reducing handling time and allowing the return of N95 respirators to their original user.

## Data Availability

All data in the manuscript are available upon request

## ACKNOWLEDGEMENTS

We would like to thank Drs. Mark Einstein, Debra Chew, Ms. Safia Amatullah, Ms. Jo Ellen Harris and Mr. Lee Clark for coordinating the N95 respirator decontamination program between UH and Rutgers and coordinating delivery and pickup of N95 respirators.

## Supplement Figure Legend

**Figure S1:**
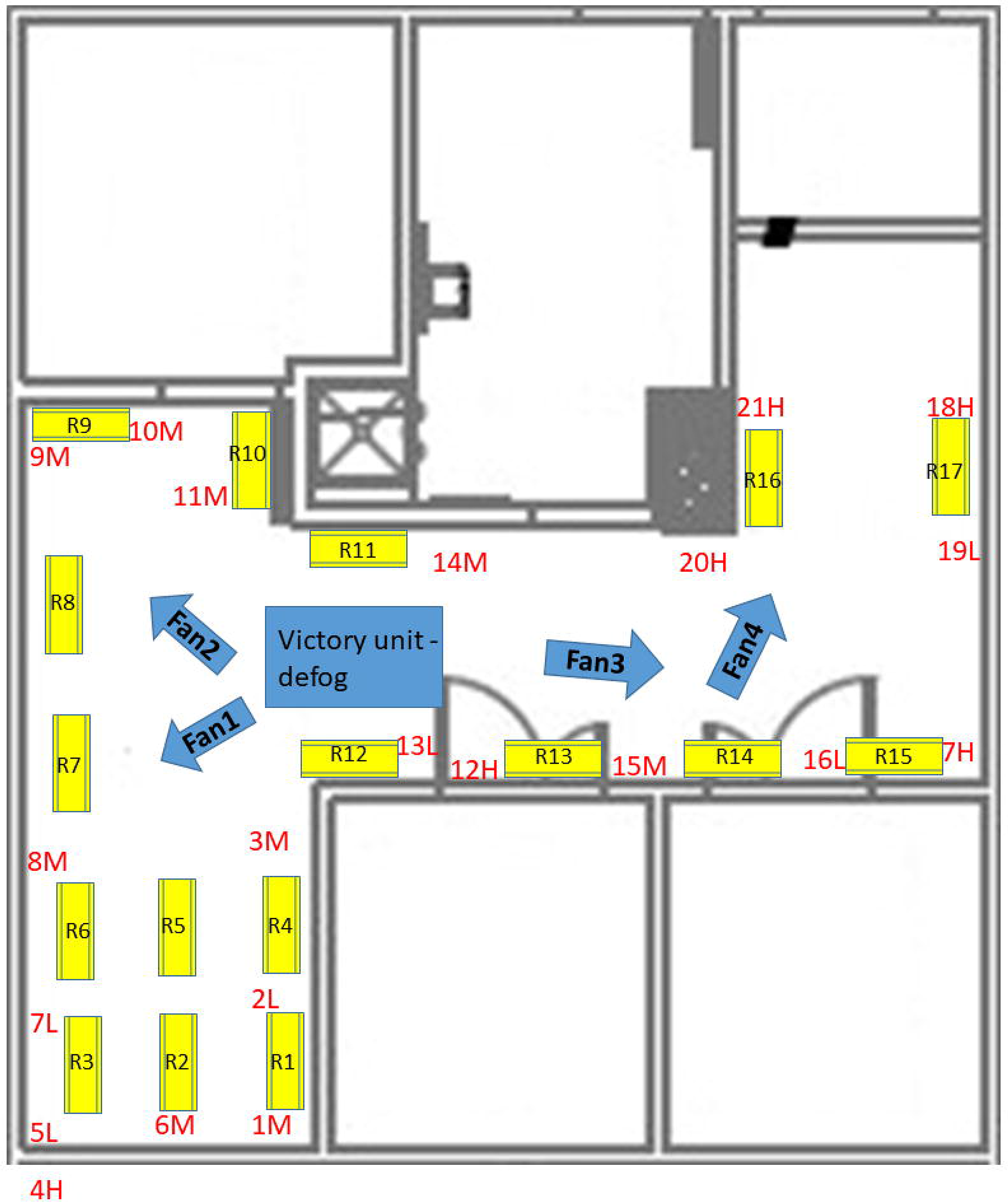
Map of the facility used for decontamination of N95 respirators. Yellow rectangles represent the locations of metal racks, blue arrows represent the location and direction of fan air flow and the blue rectangle indicates the location of the VHP Victory unit. The red numbers indicate the locations of BIs and CIs in the room and the relative position: “H” = located in the highest portion of the wall, “M” = located in the middle level of the wall, “L” = located in the lower portion of the wall toward the floor.

**Figure S2:**
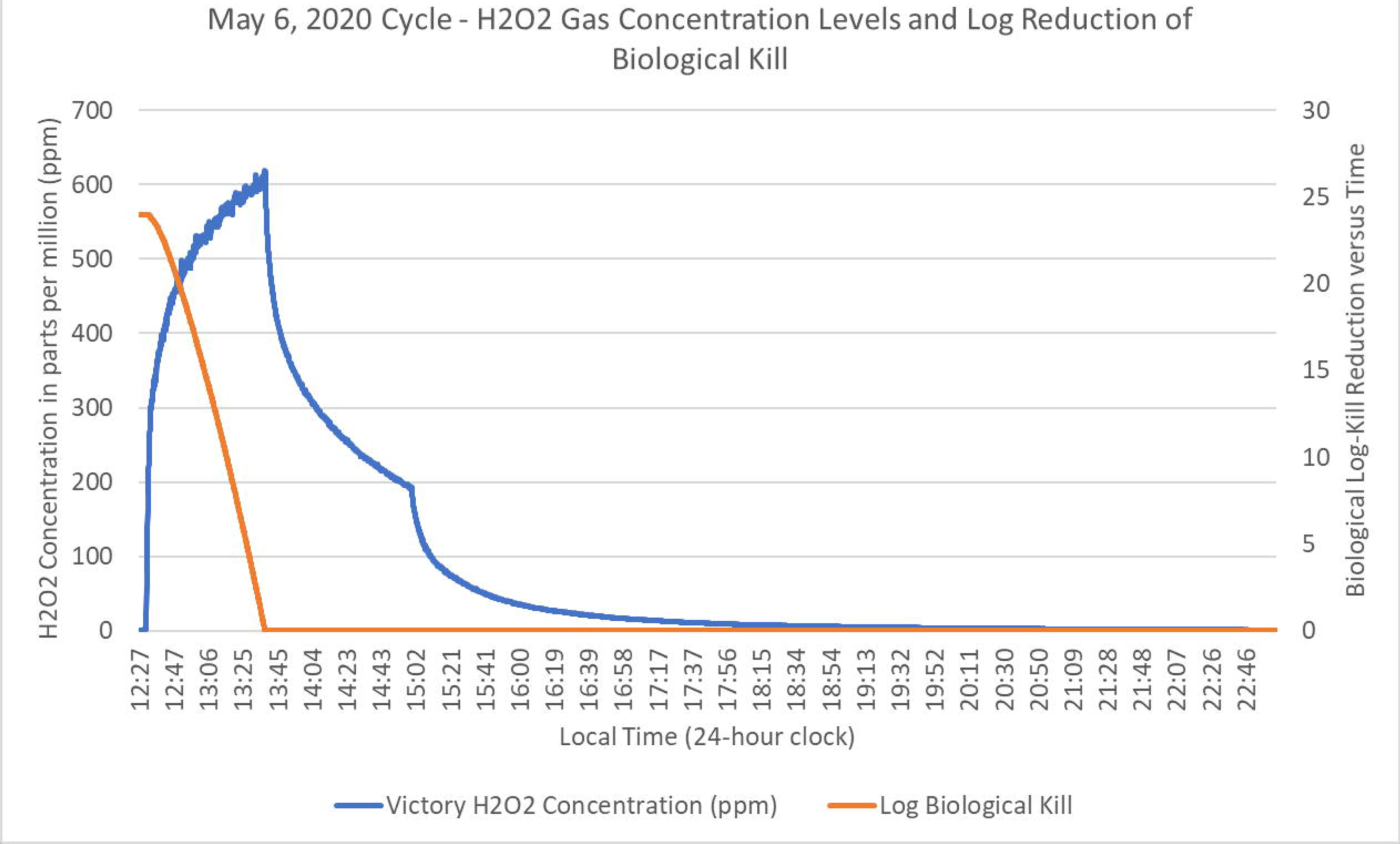
Example of a decontamination cycle. In the graph is reported the concentration of VHP as a function of time and the Log Biological Kill.

## References

[1] Dong E, Du H, Gardner L. An interactive web-based dashboard to track COVID-19 in real time. The Lancet infectious diseases 2020; 20: 533–4.

[2] Nagesh S, Chakraborty S. Saving the frontline health workforce amidst the COVID-19 crisis: Challenges and recommendations. Journal of global health 2020; 10: 010345.

[3] Tran K, Cimon K, Severn M, Pessoa-Silva CL, Conly J. Aerosol generating procedures and risk of transmission of acute respiratory infections to healthcare workers: a systematic review. PLoS One 2012; 7: e35797.

[4] Van Doremalen N, Bushmaker T, Morris DH et al. Aerosol and surface stability of SARS-CoV-2 as compared with SARS-CoV-1. New England Journal of Medicine 2020; 382: 1564–7.

[5] Herron JBT, Hay-David AGC, Gilliam AD, Brennan PA. Personal protective equipment and Covid 19-a risk to healthcare staff? The British journal of oral & maxillofacial surgery 2020; 58: 500–2.

[6] Remuzzi A, Remuzzi G. COVID-19 and Italy: what next? Lancet 2020; 395: 1225–8.

[7] O’Sullivan ED. PPE guidance for covid-19: be honest about resource shortages. Bmj 2020; 369: m1507.

[8] Whitworth J. COVID-19: a fast evolving pandemic. Trans R Soc Trop Med Hyg 2020; 114: 241–8.

[9] Fischer RJ, Morris DH, van Doremalen N et al. Assessment of N95 respirator decontamination and re-use for SARS-CoV-2. medRxiv 2020; doi 10.1101/2020.04.11.20062018.

[10] Smith JS, Hanseler H, Welle J et al. Effect of various decontamination procedures on disposable N95 mask integrity and SARS-CoV-2 infectivity. medRxiv 2020.

[11] Procedure Guidance for N95 and Facemask Reuse. In: Health ND, Ed.: 2020.

[12] Cheng VC, Wong S-C, Kwan GS, Hui W-T, Yuen K-Y. Disinfection of N95 respirators by ionized hydrogen peroxide during pandemic coronavirus disease 2019 (COVID-19) due to SARS-CoV-2. The Journal of Hospital Infection 2020.

